# Drivers of the decline in adolescent life satisfaction over 16 years: Evidence from UK representative household panel data (2009-2024)

**DOI:** 10.64898/2026.10.02.26364426

**Authors:** Niamh Dooley, Mental Health Advisory Panel, Georgina Baldwin Brown, Mary Cannon, Stephan Collishaw, Mina Fazel, Argyris Stringaris, Louise Arseneault

## Abstract

**Background:** The decline of mental health among adolescents in the UK over the past two decades has occurred amidst many societal and technological changes making it difficult to identify drivers of these trends. We aimed to identify which aspect of life showed the most rapid deterioration in satisfaction in the UK since 2009 and which aspects have become increasingly correlated with mental health.

**Methods:** Between 2009 and 2024, 15,829 adolescents aged 10-15 rated their satisfaction with life as well as five aspects of life (family, friends, school, schoolwork, appearance), via the UK Household Longitudinal Study. Population-weighted regressions were used to estimate annual summary statistics on these items (means, proportions) and the relative associations between satisfaction items and socioemotional difficulties, as measured by the Strengths and Difficulties Questionnaire (SDQ). Meta-regressions estimated linear changes over time, with interactions testing for heterogeneity across sexes, age-groups, and household income.

**Results:** Mean satisfaction with life dropped over the 16 years of study (B=-0.028, 95% CI=-0.034,-0.022), as did satisfaction with all five aspects of life. Satisfaction with school declined most rapidly (B=-0.027, 95% CI=-0.034,-0.020), becoming accelerated in the most recent 5-year window (2020-24; B=-0.139, 95% CI=-0.178,-0.099). The proportion of dissatisfied individuals rose most rapidly for schoolwork, particularly for females. Satisfaction with appearance became increasingly associated with general life satisfaction (B=0.009, 95% CI=0.005, 0.013, p<.001) and socioemotional difficulties (B=-0.051, 95% CI=-0.073,-0.029, p<.001) over time.

**Conclusions:** Our findings add to international evidence for academic pressure and school environments as plausible drivers of recent declines in life satisfaction, particularly among female adolescents. Future studies should aim to identify the specific aspects of school life that are influencing adolescent mental health and the impact of existing educational policies on student wellbeing should be evaluated.

## INTRODUCTION

Mental health problems have become increasingly prevalent among adolescents in the past few decades. Studies indicate the change has been most apparent among females, and in high-income countries (Armitage et al., 2023; Campbell et al., 2021; Castelpietra et al., 2022; Collishaw & Sellers, 2020; McGorry et al., 2024; Rudolf & Bethmann, 2022). The UK has seen significant increases in clinical diagnoses (NHS Digital, 2022; Trafford et al., 2023), parent reports of emotional problems (Armitage et al., 2023; Patalay & Gage, 2019), subjective reports of distress or dissatisfaction with life (Cosma et al., 2023; Pierce et al., 2025; Zhang et al., 2023), and deaths by suicide (Bertuccio et al., 2024) among its young people. Fifteen-year-olds in the UK reported the lowest life satisfaction levels when compared to their peers in 24 other European countries in 2018 (Marquez & Long, 2021). This wave of youth psychological distress poses a challenge for clinical services, where the demand outpaces the supply, leading to unmet need and inequalities in care provision (O’Hare et al., 2025; Udu et al., 2026). There are deep and long-term human and economic costs to untreated psychopathology in childhood (Chartier et al., 2022; Coghill et al., 2017; Copeland et al., 2021; Dooley et al., 2023; Sellers et al., 2019), so understanding the reasons for these recent but sustained population shifts in mental health are a public health priority.

Uncertainty remains around the drivers of the deterioration in mental health and life satisfaction in the UK in the 21^st^ Century. Some studies have suggested a step change occurred in the late 2000s and early 2010s (Armitage et al., 2023; Armitage et al., 2025; Zhang et al., 2023), predating the COVID-19 pandemic, and aligning with a range of societal changes including financial austerity and inequality following the global recession (Brown et al., 2024; Collishaw et al., 2019; Viner et al., 2017), greater awareness and destigmatisation of mental illness (Foulkes & Andrews, 2023; Marcotulli et al., 2026) and increased use of digital technology (Twenge, 2020). However, the overlapping timelines of these changes makes it difficult to disentangle their effects. Most studies comparing plausible theories for the deterioration in adolescent mental health have been reviews (Armitage et al., 2024; Pierce et al., 2025), in which the strength of evidence for each theory can be conflated with the quantity or quality of evidence available. There is therefore a need for a comparison of plausible explanations in a single nationally representative study.

In this study, we investigated changes in young peoples’ satisfaction with various aspects of their lives to identify drivers of mental health deterioration over the past 16 years. Life satisfaction is a subjective appraisal of one’s life and is considered to be one aspect of general wellbeing (Metler & Busseri, 2017; Steptoe, 2019), independent from positive and negative affect, and purpose in life. It is related to psychopathology, with longitudinal studies demonstrating reciprocal associations with psychiatric diagnoses (Fergusson et al., 2015; Lombardo et al., 2018; Myers & Diener, 1995). We take a social systems approach to the question of why adolescent wellbeing and mental health has deteriorated in the UK, examining how young people’s satisfaction levels have changed with core social systems including home, school and peer life. Most of childhood is spent within these social networks and some of the strongest childhood risk factors for mental disorder (e.g., parental maltreatment, peer victimisation) fall within these social domains (Arseneault et al., 2008; Baldwin et al., 2023; Moore et al., 2017). In considering school life, we distinguish between a child’s satisfaction with *schoolwork* and their satisfaction with *their school*. Evidence indicates that rising academic pressure in recent years may have contributed to increases in mental health symptoms, with a greater perceived academic pressure in females compared to males, consistent with the sex difference in mental health reports (Armitage et al., 2025; De Looze et al., 2025; Rudolf & Bethmann, 2022; Steare et al., 2023). However, growing dissatisfaction with broader aspects of school life such as belonging and support from teachers has also been observed in England (Academy of Medical Sciences & British Academy, 2025), making it difficult to identify whether academic pressure or the wider school environment is driving the change. Many believe that increased exposure to social media has amplified adolescents’ feelings of inadequacy, specifically with their appearance. Relevantly, the incidence of eating disorder presentations to primary care clinics in the UK increased significantly between the 2000s and the 2020s, specifically in adolescent girls (Trafford et al., 2023). This trend, along with the association between low self-image and subsequent psychopathology (Creese et al., 2023), supports the hypothesis that rising dissatisfaction with appearance (or strengthening association with mental health) may underlie deteriorating mental health in adolescents.

In this study, we first examined which aspects of adolescent life showed the most rapid declines in satisfaction over the past 16 years. Second, we identified which aspects of life had changing associations with a single-item measure of life satisfaction and a multidimensional measure of socioemotional difficulties. We tested this as it may be that the impact of risk factors on mental health has changed rather than their prevalence. Third, we explored whether changes over time differed by sex (males, females), age group (10-12, 13-15 years old), or household income (low, middle, high).

## MATERIALS AND METHODS

### Data source

The UK Household Longitudinal study (UKHLS), also known as Understanding Society, is a longitudinal study tracking approximately 40,000 households across England, Scotland, Wales and Northern Ireland since 2009 (ISER, 2023). Participants are followed annually unless all members of a household refused further participation, withdrew consent, emigrated, died, or became unable to give informed consent. New members joining a household (e.g., partners or children) are included if they reside with a core sample member, and core sample members are followed to other dwellings. At the time of analysis, 15 waves of data collection had been completed across 16 calendar years. Households are followed up in the same quarter every year (ISER, 2023).

All 10-15-year-olds in participating households were invited to complete a paper-based questionnaire, with permission from their parent/carer. When adult interviews were held in-person, the young person completed the survey and returned it to the study team on the same day. If adult interviews were completed over the phone, the young person was mailed the survey and asked to post it back. An unconditional £5 incentive accompanied the youth survey.

The scheduled date of household interview was used as the estimated time of youth survey completion which may have differed from the actual youth survey completion due to scheduling difficulties and postal surveys. However, 97% of interviews took place within 3-months of the scheduled date.

Ethical approval was received from the University of Essex Ethics Committee at each study sweep (ISER, 2023). Adult participants provided informed oral consent for their participation in UKHLS before the face-to-face or phone interview.

### UK population representativeness

The UKHLS employed a stratified, clustered, probability sampling design to ensure representativeness of individuals living in private households in the UK in 2009. The sample was boosted in 2014 and again in 2022 to ensure new and pre-existing minority groups were represented. UKHLS does not include young people without stable housing, in care or other institutions.

Cross-sectional youth weights were used in all analyses which adjust for: (1) unequal selection probability in UKHLS design; (2) wave 1 non-response; (3) attrition, and (4) within-household non-response to the youth survey. Both the study design and weights were intended to make estimates representative of the UK population however non-response related to health or wellbeing may not be fully accounted for e.g. attrition in ethnic minority groups (Cabrera-Àlvarez & Lynn, 2023). We analysed the demographic shifts for participants over time and performed non-response analyses to explore how non-response may have influenced results (Figures S9, S10).

## Measures

### Life satisfaction

Adolescents were asked to rate their feelings about aspects of their lives. Responses were provided on a 7-point visual scale ranging from a smiling to a sad face (Figure 1, top). Participants were instructed to: *“tick the box which comes closest to expressing how you feel about…your schoolwork…your appearance…your family…your friends…the school you go to*…*your life as a whole”.* Each item was treated as a separate construct given our interest on inter-correlations between items. Pearson’s correlations between satisfaction items ranged between.20 and.60 (Figure S1). We recoded responses on a scale from +3 (most happy) to –3 (most sad) with 0 corresponding to a neutral face. These variables were treated as continuous, based on simulations showing variables with more than five categories are approximately continuous (Rhemtulla et al., 2012). We also dichotomised the scale to calculate the proportion of participants reporting dissatisfaction in each domain (rating<0). The construct validity of single-item subjective life satisfaction is supported by concurrent and prospective correlation with other wellbeing and mental health indicators (Diener et al., 2018; Spinhoven et al., 2021).

**Figure 1.**
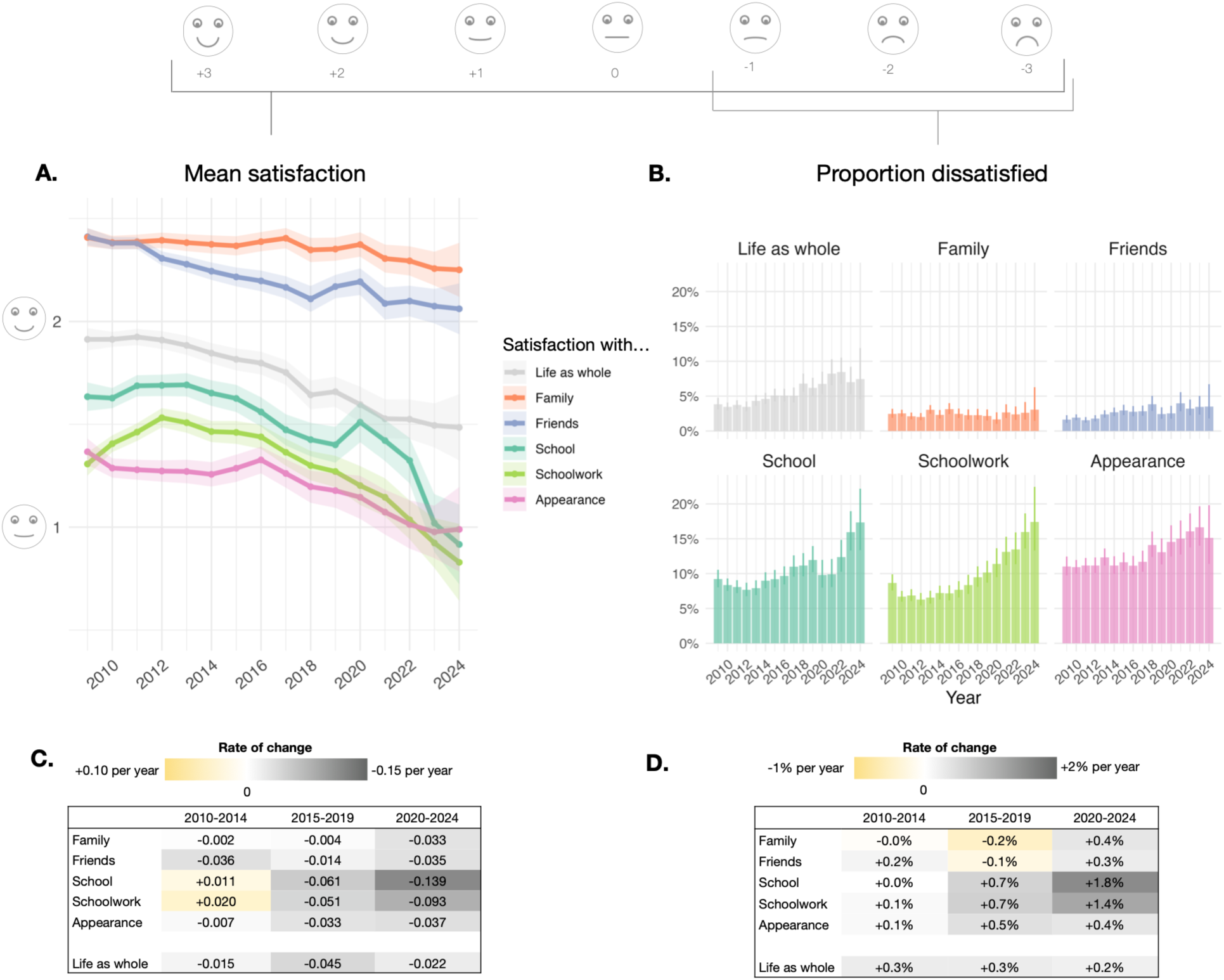
**A.**Population-weighted cross-sectional mean satisfaction ratings (error band = 95% confidence interval). **B.** Population-weighted cross-sectional proportion of unhappy individuals (error bar = 95% confidence interval). C. Change statistics within 5-year windows for mean satisfaction ratings D. Change statistics within 5-year windows for proportion of unhappy individuals.

### Socioemotional difficulties

Socioemotional difficulties were assessed using the Strengths and Difficulties Questionnaire (SDQ; (Goodman, 1997) which consists of 25 items with responses “Not True” (0) to “Somewhat True” (1) or “Certainly True” (2). Twenty items were combined into a total difficulties scale (range 0-40) which captures difficulties in attention/hyperactivity, emotional, peer relationship, and conduct problems. The SDQ was self-completed by adolescents in every second wave of UKHLS, leading to a restricted sample. Including socioemotional difficulties in this analysis allows extension of findings from a single-item measure of life satisfaction to a clinically-validated scale of mental health problems (Vugteveen et al., 2021), and triangulation across two constructs relevant to mental health (Steptoe, 2019). The SDQ is also a continuous multi-item measure providing greater variance and statistical power than the single item Likert scale.

### Demographics

Adolescents were asked “Are you male or female?” at each wave. An adult in the household also reported the sex of these young participants as part of the household grid. If sex reporting was inconsistent across waves or reporters, it was coded as “inconsistent” by UKHLS and as missing in the current study (n<0.1%; Table 1). Household income was captured by the equivalised net income per month, i.e., the sum of monthly total net personal income (no deductions) received by all household members, divided by the number of adult equivalents in the household. The number of household members was determined by an equivalence scale, which assigns a weight of 1 to the first adult (person aged 14 or older), a weight of 0.5 to each additional adult, and a weight of 0.3 to each person aged 0-13. Tertiles were then created for each year such that evenly sized low-, middle-and high-income groups were based on the equivalised incomes of that year. Due to repeated observations of the same household with multiple sources, the UKHLS can compute derived demographic data where it is unavailable or inconsistent therefore there is no or very little missing data on age (n=2), household income (n=0), and ethnicity (n=94).

**Table 1.** Characteristics of UKHLS analytic sample.

| Sample characteristic |  |
| --- | --- |
| **Number of participants** | 15,829 (with 45,814 observations) |
| **Sex, n (%)** |  |
| Male | 7921 (50.0%) |
| Female | 7901 (49.9%) |
| Inconsistent | 7 (<0.1%) |
| **Age, in years<sup>1</sup>** |  |
| Mean (SD) | 11.5 (4.96) |
| Min-Max | 10 – 15 |
| **Year of birth** |  |
| Mean (SD) | 2001 (4.96) |
| Min-Max | 1993 – 2013 |
| **Number of observations per participant, n (%)** |  |
| Mean (SD) | 2.8 (1.67) |
| Min-Max <sup>2</sup> | 1 – 7 |
| **Country, n (%)<sup>1</sup>** |  |
| England | 12,788 (80.3%) |
| Scotland | 1,251 (7.9%) |
| Wales | 897 (5.6%) |
| Northern Ireland | 978 (6.1%) |
| **Ethnicity, n (%)** |  |
| British or Irish | 10,961 (70.1%) |
| Any other white background | 315 (2.0%) |
| South or East Asian | 2,196 (14.0%) |
| African or Caribbean | 934 (6.0%) |
| Mixed background | 827 (5.3%) |
| Other | 410 (2.6%) |
| **Number of households** | 9,341 |
| **Number of respondents (aged 10-15) from same household** |  |
| Mean (SD) | 1.69 (0.91) |
| Min-Max | 1 – 12 |
| **Household net monthly income** |  |
| Mean (SD) | £3,335 (£2,447) |
| Min-Max <sup>3</sup> | -£1,391 - £55,200 |

<sup>1</sup> Age and country of residence at point of entry to the study.
<sup>2</sup> A small number of participants (n=53) completed the youth survey 7 times instead of the expected max of 6.
<sup>3</sup> Self-employed individuals could report negative net labour income.

### Pre-registration

The study protocol was pre-registered on Open Science Framework (https://osf.io/3sc5b), with any deviations detailed in the version history.

### Public involvement

A group of young people aged 16 to 27 with lived experience of mental health difficulties in their adolescence aided with the design and interpretation of this secondary data analysis. The group was recruited via mental health charities in the UK (McPin Foundation) and Ireland (spunout). A discussion of the planned research questions (17/09/2025), methods and results (21/02/2026) of the study took place in online focus groups (90 minutes each). Attendance ranged from 7-11 members at each meeting. Input from the group has been included in the results and discussion, paraphrased in collaboration with members.

## Statistical analysis

We estimated cross-sectional population averages in means and proportions (and their standard errors) for each calendar year using intercept-only general linear models, incorporating cross-sectional weights and accounting for the clustered stratified sampling design.

To estimate changes over time, we entered cross-sectional estimates into a meta-regression with time as a numeric predictor (0=2009). A variance-covariance matrix was included in this model, which accounted for the repeated measurement of the same households over time (Table S1), and the variance for the point estimate for each year. Two variants of the meta-regression estimated the linear change (i) over all available 16 years (2009-2024), and (ii) within 5-year windows (2010-2014/ 2015-2019/ 2020-2024). We excluded 2009 from the latter analysis to create three equally sized time windows as it was the earliest observation. As a sensitivity analysis, we excluded the years of the COVID-19 pandemic and associated lockdowns (2020-2021) and re-ran linear change analyses across the full period.

We assessed the relative influence of satisfaction in sub-domains on general life satisfaction using repeated cross-sectional weighted regressions. For each year, a model estimated the relative association between satisfaction in each domain (univariately & adjusting for all other domains) and general life satisfaction. We extracted cross-sectional weighted beta coefficients and their standard errors for each sub-domain for each year. Changes over time in these correlation estimates were assessed using a meta-regression with time as a predictor, and the same model parameters as described above. We repeated the analyses using socioemotional difficulties instead of life satisfaction to assess the measure and construct-specificity of the findings. We explored differences across sex, age, and household income by including these variables in the meta-regression with an interaction term between group and time.

For inferential analyses that test linear trends over time, we applied a Bonferroni-corrected p-threshold that accounts for number of outcomes tested (e.g., *p*=.05/6 domains of satisfaction=.008). Case-wise deletion (within-year) was used if participants were missing data on any variable. The analytic sample size ranged from 5199 (in 2010) to 728 (in 2024; Table S2).

All R code used for data manipulation and analyses are publicly available (https://rpubs.com/dooleyr). Further details on the statistical analysis are available in the Supplemental Methods.

## RESULTS

### Declining life satisfaction over time

We observed a decline in mean levels of life satisfaction and an incline in socioemotional difficulties over the 16-year period of this study, from 2009 to 2024 (Figure 1A; Figure S7). Satisfaction in all five aspects of life also decreased over this period (Figure 1A), with the steepest decline in satisfaction with school, followed by friends, appearance, and schoolwork (Table 2). The decline in satisfaction with family was not significant at the Bonferroni-corrected p-threshold. Despite decreases over time, all mean satisfaction estimates remained in the “positive” range (i.e., smiling faces) throughout the study period.

**Table 2.**
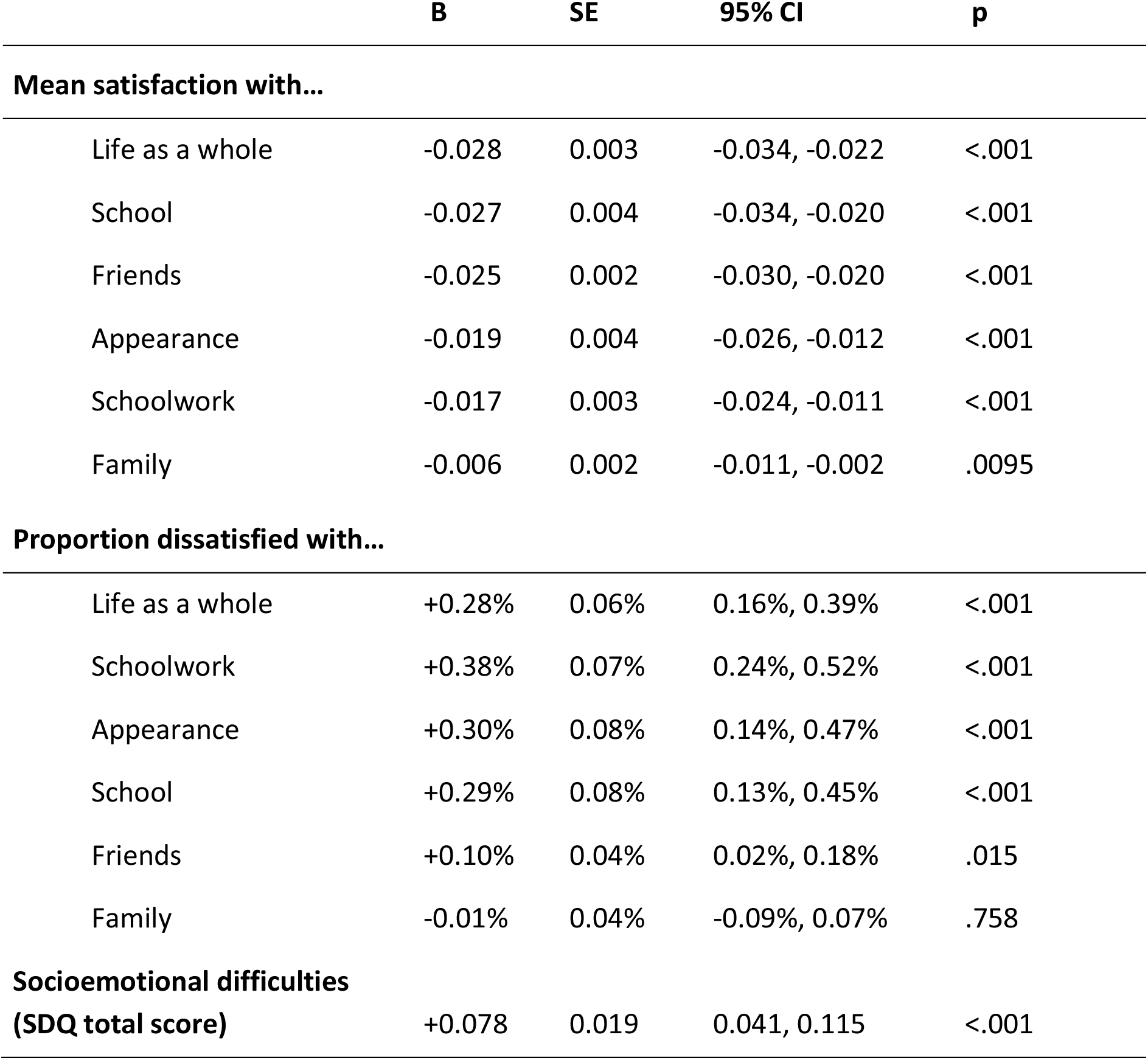
Linear slope estimates (B) reflecting the annual rate of change in satisfaction, dissatisfaction, and socioemotional difficulties, averaged across 16 years (2009-2024)

The proportion of individuals dissatisfied with their life (selecting “sad” faces on the visual response scale) nearly doubled in the most recent 5-year window compared to the first 5 years (from 3.9% in 2010-2014 to 7.6% in 2020-2024; Figure 1B). Over the full study period, dissatisfaction with schoolwork had the steepest rise, followed by appearance and school (Table 2). The proportion of individuals dissatisfied with their friends and family did not significantly change. Estimates for schoolwork, appearance and school indicate that the proportion of dissatisfied individuals increased by less than 0.5% each year, but 4-6% over the 16-year period.

Breaking the study period into 5-year windows showed that decreases in mean satisfaction, and increases in the proportion of dissatisfied individuals, accelerated in the most recent time window between 2020 and 2024 (Figure 1C-D). The life domains with the most rapid decline were school and schoolwork, though the decline can be observed from 2015 onward (Figure 1C). Pairwise z-tests showed that downward slope in satisfaction with school and schoolwork became progressively steeper between the first and second windows (zsch_ool_=4.16, *p*<.001; z_schoolwork_=4.08, *p*<.001), and between the second and third windows (z_school_=5.85, p<.001; z_schoolwork_=4.32, *p*<.001). The change in satisfaction with life as a whole also accelerated significantly between the first and second windows (z_school_=3.67, p<.001) but did not change significantly in the third 5-year window.

We investigated whether changes happened at a similar rate across sexes, ages and household income groups. While we report average group differences across all 16 years (main effect of group only), our focus was on the widening/narrowing of these differences over time (interaction of time and group). Significant sex differences were observed in satisfaction with schoolwork (male<female), appearance (female<male) and life as a whole (female<male). There was also a sex difference in the rate of change of satisfaction with friends, schoolwork, and life such that satisfaction declined more rapidly over the study period in females compared to males (Interaction B_friends_=0.017, 95% CI=0.008, 0.026, *p*<0.001; B_schoolwork_=0.019, 95% CI=0.007, 0.031, *p*=0.003; B_life_=0.024, 95% CI=0.013, 0.036, *p*<0.001). The rate of change in appearance satisfaction was not significantly different in males compared to females, however sex-specific trends in this domain were non-linear. Adolescents aged 13-15 reported lower satisfaction levels in all areas of life compared to the younger group (aged 10-12) but only showed a significantly more rapid decline in satisfaction with friends and life as a whole (Interaction B_friends_=0.015, 95% CI=0.006, 0.024, *p*=.001; B_life_=0.018, 95% CI=0.007, 0.029, *p*=.002). Adolescents from high income homes rated their satisfaction with family, school, schoolwork and life in general significantly higher than those from middle-income homes. However, the rate of change of satisfaction over time was not significantly different across income groups, for any aspect of life (Figure 2).

**Figure 2.**
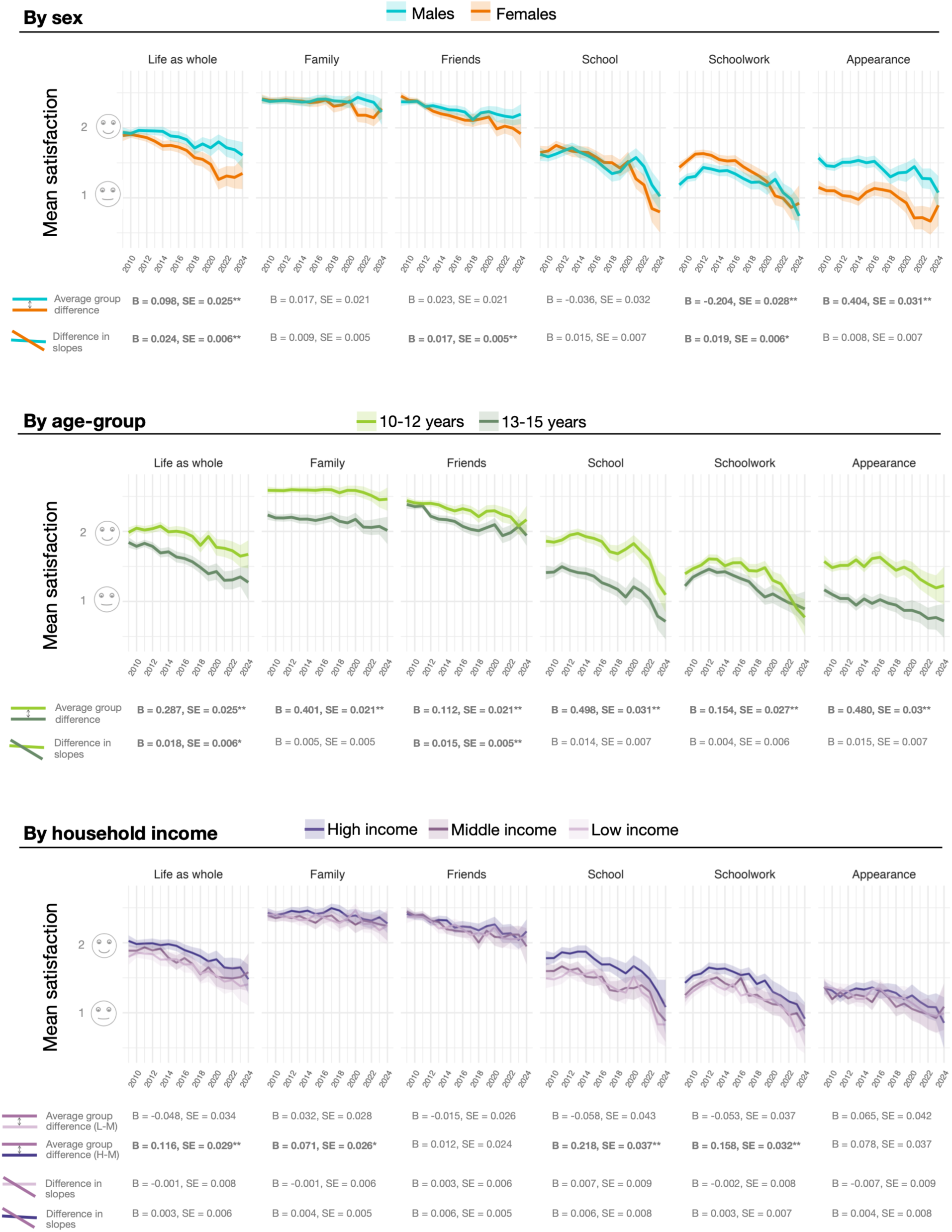
Trends in life satisfaction across 2009-2024 split by sex, age-group and household income

**Figure 3.**
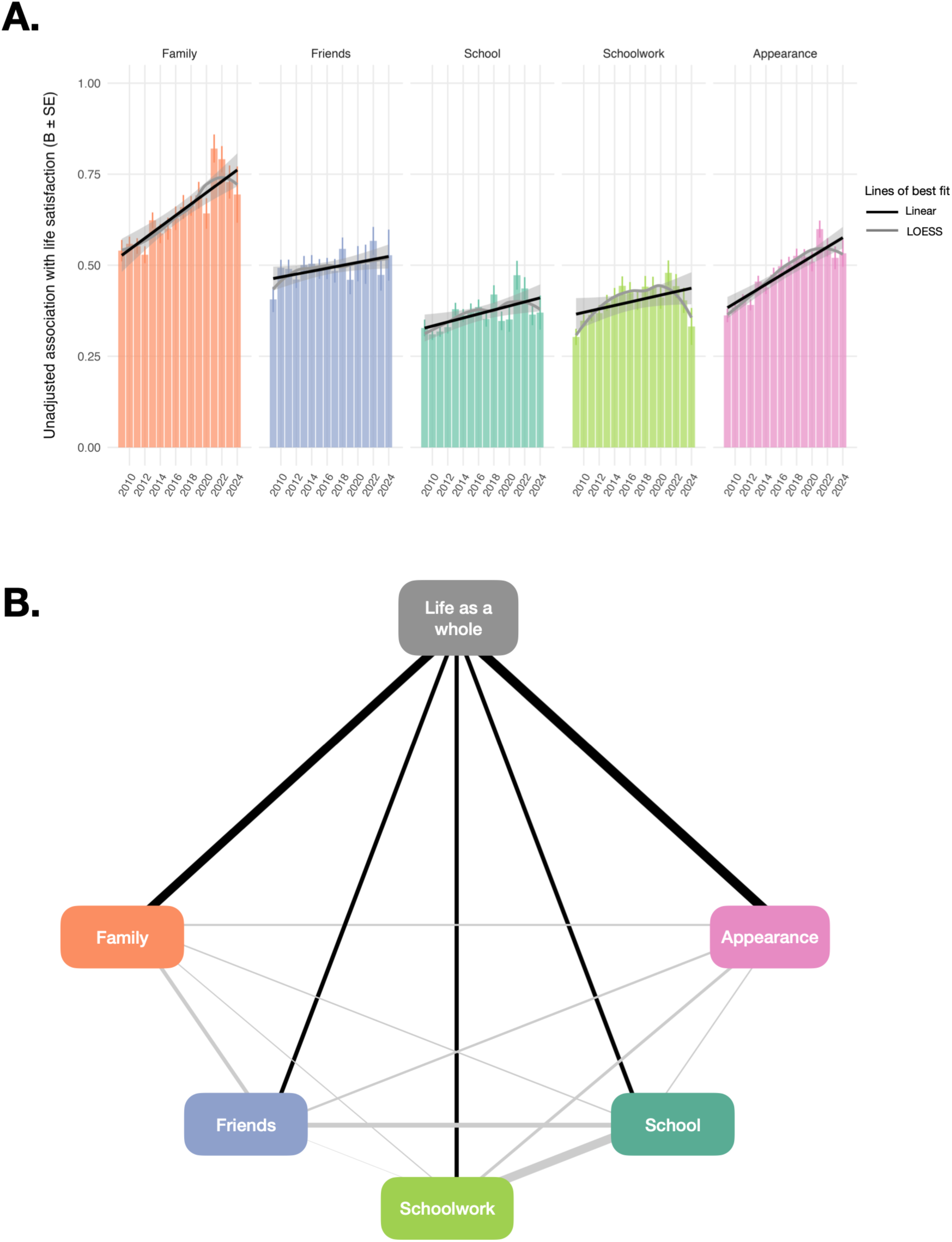
**A.** Changes in associational strength over time between satisfaction in five domains of life and general life satisfaction across the study period. **B.** Network diagram showing partial correlations between satisfaction with life and five sub-domains, across all years.

Several sensitivity analyses were conducted to test robustness of findings. First, we confirmed that the linear decline in satisfaction levels over 16 years remained significant when years of COVID-19 pandemic were removed (i.e., 2020-2021; Tables S3, S4). Second, we tested whether the population variance in satisfaction was increasing over time (in addition to a declining mean) which would suggest growing heterogeneity within the population. Variance grew significantly over the study period for satisfaction in all domains, except school and family (Table S6; Figure S6). However, distributions of satisfaction remain unimodal at all years indicating shift of the unimodal distribution rather than a polarisation of responses (Figure S2). Third, based on input from the lived experience group, we created an alternative three-way age grouping as a more nuanced measure of developmental stage (10-11/12-13/14-15 years). Changes in satisfaction over time were not significantly different between 12-13-year-olds and 14-15-year-olds (Figure S5).

### Relative influence of domains of life on general life satisfaction

We assessed the associations between satisfaction with each aspect of life and general life satisfaction. In univariate models, life satisfaction was most strongly correlated with satisfaction with family, followed by friends, appearance, schoolwork and school (B_family_=0.62, 95% CI=0.59-0.64; B_friends_=0.49, 95% CI=0.47-0.52; B_appearance_=0.45, 95% CI=0.43-0.47; B_schoolwork_=0.38, 95% CI=0.35-0.40; B_school_=0.34, 95% CI=0.32-0.36). In multivariate models adjusting for all other aspects of life, satisfaction with family remained most strongly associated with life satisfaction (B=0.33) and satisfaction with school the least (B=0.10; Table S5).

We observed a significant increase in the association between appearance and life satisfaction (B=0.014, 95% CI=0.011 – 0.018, *p*<.001), even after adjusting for satisfaction in other domains (B=0.009, 95% CI=0.005-0.012, *p*<.001). When replacing life satisfaction with a measure of socioemotional difficulties, we found a similar pattern whereby the magnitude of the association between satisfaction with appearance and socioemotional difficulties grew significantly over time (B=-0.051, 95% CI=-0.073,-0.029, *p*<.001; Figure S8). Associations also strengthened between satisfaction with family and life as a whole (Table S5); and between satisfaction with family and socioemotional difficulties (B=-0.049, 95% CI=-0.081,-0.017, *p*=.003).

Regarding differences across demographic subgroups, the association between satisfaction with family and life satisfaction was lower in males compared to females (group difference B_adj_=-0.070, 95% CI=-0.12,-0.02, *p*=.006). However, we did not find significant group differences in the *change in association* over time for any domain. The absence of a group-by-time interaction on the association between appearance and general life satisfaction indicates that the association increased at a similar rate in all sex, age and income groups.

## DISCUSSION

This nationally representative study of over 15,000 adolescents in the UK comes at a crucial time, amid growing concerns over youth mental health. We observed that on average, adolescents in the UK are satisfied with their lives. However, we observed a relatively steady rise in both dissatisfaction with life and socioemotional problems over the 16-year study period. School was identified as a plausible driver of these population shifts in global wellbeing measures, as the aspect of life with the most rapidly declining satisfaction levels. Schoolwork was also implicated, characterised by the most rapidly growing rates of dissatisfaction. These two related aspects of life emphasise the need to understand which specific aspects of academic life are influencing adolescent mental health. We found that satisfaction with appearance was increasingly correlated with socioemotional difficulties and general life satisfaction over time, which may be linked to increased time spent on image-based apps

Could growing dissatisfaction with school and schoolwork be a driver of mental health deterioration among adolescents in the UK since the mid-2010s? First, in support of this, we found dissatisfaction with school and schoolwork rose while dissatisfaction with family and friends did not, demonstrating domain specificity in our findings. Second, the drop in satisfaction with schoolwork was steeper for females compared to males, which is consistent with the sex difference in mental health trends (Armitage et al., 2023; Campbell et al., 2021). And third, our findings add to a rapidly growing evidence base demonstrating the contribution of academic pressure to mental health deterioration among adolescents in high-income countries over the past two decades (Armitage et al., 2025; Boer et al., 2023; Cosma et al., 2020; De Looze et al., 2020; Guo et al., 2026; Rudolf & Bethmann, 2022). That said, a systematic review reports that causal evidence is lacking (Steare et al., 2023). It remains possible that an unmeasured variable (e.g., generalised anxiety, screen use, sleep deficits) influences both satisfaction with school and general wellbeing independently, or that there is reverse causality. Our youth lived experience group resonated with the deterioration in school satisfaction after COVID-19 and mentioned a deterioration of in-person social skills, a widening of educational inequalities during remote learning, and the challenge of re-adjusting to a highly structured environment. Schools play a central role in adolescent mental health support, but our findings indicate that the school may also be a source of discontent. There is a need to identify the modifiable school-based factors that are influencing mental health on a population level including but not limited to, curriculum and assessment policies (Cashman et al., 2023), preparedness for secondary school (Donaldson et al., 2022), and feelings of belonging and connectedness in school (Raniti et al., 2022).

Historically, appearance was the aspect of life with the lowest average satisfaction levels among UK adolescents. Also concerning was the growing association between satisfaction with appearance and broad measures of wellbeing like socioemotional difficulties and satisfaction with life in general. This trend was not driven by one sex, age-group, or income bracket more than another, implying a growing influence of self-image to wellbeing on a population level. The youth lived experience panel was not surprised at this result and believed that young people were more insecure in their appearance now compared to 10 years ago. It is possible that the rising popularity of image-based platforms such as Instagram (since 2013) and TikTok (since 2017) has led to an increased awareness or societal emphasis on appearance. Longitudinal and experimental studies have demonstrated that exposure to imagery encouraging social comparison can have a harmful effect on adolescent mental health, via low body image and disordered eating (de Valle et al., 2021; Faelens et al., 2021; Fioravanti et al., 2022; Holland & Tiggemann, 2016). Early adolescence (11-15-years) is also a developmentally sensitive window in which social media may have a more pronounced effect on subsequent wellbeing (Orben et al., 2022). While our study did not directly measure use of digital media, our finding of rising dissatisfaction with appearance, particularly from around 2017, points to digital media use as a plausible factor. Proposed policies in the UK aimed at delaying access to social media and restricting content (Woodhouse et al., 2026) should be evaluated in terms of its effect on adolescents’ satisfaction with their appearance as one possible mechanism to improving general mental health.

The drop in the mean population satisfaction with life could be considered small, which raises the question of how clinically relevant this finding is. It is important from a public mental health perspective because our results show the entire distribution of life satisfaction has shifted in this population, the drop in mean being just one feature of this (Figure S2-3). In public health, the case is often made that a small change to the population mean of a risk factor of disease has a greater influence on the prevalence of that disease, than a large charge in the group of individuals with a high risk of the disease (Carey et al., 2023; Rose, 1985). A shift in symptom distribution is also important for clinicians, as it would help explain the increase in extreme cases (e.g., diagnosis of mental disorder in adolescents (Collishaw & Sellers, 2020; McGorry et al., 2024; O’Hare et al., 2025)) but also the increase in referrals and cases around the threshold for diagnosis. Our findings align with other evidence (Fazel & Soneson, 2023; Lo Moro et al., 2020) supporting the use of universal or population-level preventative interventions to “shift the curve” and we point to several domains of life in which these interventions may be most relevant.

A powerful feature of repeating the same survey over 16 years in a nationally representative sample is the ability to distinguish stable group differences from changing ones. For instance, females reported lower life satisfaction than males in all years (consistent with other countries (Campbell et al., 2021)) but this sex difference widened in 2018 and was at its widest in 2021. Females aged 13-15 in particular were characterised by low satisfaction with their life and appearance, the latter showing a continuous decline since 2016 (Figure S4). Our study also adds to the accumulating evidence for academic pressure as a contributor toward the gender gap in mental health symptoms (De Looze et al., 2025; Hogberg et al., 2020; Rudolf & Bethmann, 2022; Steare et al., 2023) and suggests that satisfaction with friends may be another emerging area of difference between males and females. Those aged 13-15 displayed more concerning trends than their younger counterparts, with more rapid drops in satisfaction with life and friends over the 16-year study period. However, 10-12-year-olds displayed rapidly deteriorating satisfaction with schoolwork in the most recent 5 years. This could be explained by COVID-related delays in reading, writing and maths during primary school (Milanovic et al., 2023; Weidmann et al., 2021), compounding on entry to secondary school. Finally, school and schoolwork satisfaction differed across household income groups (high>low-middle income) which is consistent with evidence that there are economic inequalities in enjoyment and performance at school (Horanicova et al., 2022; Song et al., 2025) but our results indicate these inequalities have not widened. This is consistent with findings from repeated cross-sectional surveys of secondary school students in Wales, where schoolwork pressure rose at a similar rate, regardless of family affluence, from 2009 to 2022 (Armitage et al., 2025).

Our study has limitations. First, the cross-sectional nature of our analysis means reverse causality cannot be ruled out. For instance, it may be that poor mental health influences dissatisfaction with appearance rather than the reverse. However, even if the relationship is in the other causal direction, or bidirectional, its strengthening over time remains important information. Second, additional analysis showed that adolescents with lower life satisfaction were more likely to drop out of the study indicating sampling bias. However, if our sample increasingly under-represents adolescents with low life satisfaction over time, then our finding of declining life satisfaction over 16 years is likely a conservative estimate of the true population decline. Other predictors of non-response included ethnicity, age, country, and household income (Figure S10), which may only partially be accounted for by weighting. Third, the domains of life included in this study are not exhaustive and there are likely unmeasured confounds. However, the five domains included are pragmatic in that they capture the broad social networks and common areas of concern in adolescent life, which is invaluable in guiding further research. Fourth, one’s rating of satisfaction may be influenced by transient events and change rapidly. That said, we would expect noise due to recent personal circumstances to be similarly distributed across all years, therefore it should not affect our interpretation of temporal trends.

## Conclusion

Adolescents in the UK in the early 2020s were significantly less satisfied with their lives compared to adolescents in the early 2010s. Rising dissatisfaction with school and schoolwork suggests that education settings should be viewed not only as settings in which mental health problems emerge but as central platforms for prevention. Schools are uniquely positioned to test and deliver interventions at scale, with change to national educational policy, or at the school level. The growing association between appearance satisfaction and mental health highlights the need for early support around body image and social comparison. Causal and interventional research is needed to inform which population-level prevention strategies are effective, and for whom.

## Ethical information

Ethics approval was received from the University of Essex Ethics Committee at each wave of survey (for details see Institute for Social and Economic Research, 2023). Adult participants provided informed oral consent for their participation and their child’s participation. For further information on consent processes, see https://www.understandingsociety.ac.uk/documentation/mainstage/consents/. As this secondary data analysis aligned with the UK Data Service original consent and raised no ethical issues, it did not require additional ethics approval.

## Contribution statement

ND and LA conceptualised and designed the study. ND performed analyses and drafted the manuscript. SC, AS, and MF advised on the methodology and interpretation. The Mental Health Advisory Panel provided additional insights on the findings based on their lived experience. All authors critically reviewed and approved the final manuscript.

## Open access

For the purposes of open access, the author has applied a Creative Commons Attribution (CC BY) licence to any Accepted Author Manuscript version arising from this submission.

## Data availability

The data used in this study are available in a public, open access repository, the UK Data Service (doi:10.5255/UKDA-SN-6614-15).

